# Mechanistic Multi-Task Logistic Regression: An Alternative to Parametric Hazard Models in Pharmacometric Joint Time-to-Event Analysis

**DOI:** 10.64898/2026.08.15.26360512

**Authors:** Kuteesa R. Bisaso, Karyaburo R. Kadada, Karungi S. Bisaso, Ene I. Ette

**Author notes:** Corresponding Author: Kuteesa Ronald Bisaso.

## Abstract

**Background:** Parametric time-to-event models require specification of a baseline hazard function, which may influence prediction when the underlying hazard shape is uncertain. This study compared conventional joint longitudinal–time-to-event models with mechanistic Multi-Task Logistic Regression, which directly models the survival distribution without selecting a parametric hazard family.

**Methods:** Two complementary analyses were conducted. First, a simulated dataset of 100 individuals with longitudinal sum of longest diameters and event outcomes was analyzed using a shared mechanistic tumour shrinkage-regrowth model. Second, the same event-model families were applied to a clinical progression-free survival dataset containing 453 patients and 1,346 longitudinal SLD observations. Event submodels comprised exponential, Gompertz, Weibull, log- normal, log-logistic, and periodic or circadian hazards, mechanistic MTLR, and a hybrid neural- mechanistic extension. The simulated analysis used five-fold cross-validation, dynamic discrimination, Brier scores, integrated Brier score, calibration, and event-interval log score. The clinical case study used joint-estimation diagnostics and model-specific simulation-based longitudinal and PFS visual predictive checks.

**Results:** In the simulated dataset, longitudinal parameter estimates were comparable across models. The log-normal hazard achieved the lowest overall integrated Brier score (0.1928), whereas mechanistic MTLR achieved the highest later-landmark discrimination (AUC 0.867 versus 0.798 for all hazard models) and the lowest mean event-interval negative log score (2.362). In the clinical dataset, all eight models met numerical convergence criteria. The SLD-event association was positive across all event formulations. The periodic hazard had the lowest AIC among continuous-time hazard models but estimated a period of approximately 58 days, consistent with scheduled progression assessment. Mechanistic and hybrid MTLR showed the strongest descriptive PFS VPC calibration, with 93.4% and 95.9% coverage of the observed Kaplan-Meier curve, respectively. The hybrid nonlinear weight was small and imprecise.

**Conclusions:** The simulated and clinical analyses jointly support mechanistic MTLR as a practical complementary approach to conventional joint hazard models. Parametric hazards can provide strong probabilistic accuracy when the hazard family is well chosen, whereas mechanistic MTLR avoids continuous baseline hazard-family selection and can provide competitive discrimination and event-time distribution prediction.

## Introduction

Time-to-event (TTE) modeling is an important component of pharmacometrics for characterizing the timing of clinically meaningful events such as death, disease progression, relapse, treatment discontinuation, or toxicity . Unlike analyses that consider only whether an event occurred, TTE methods incorporate when it occurred, accommodate censoring, and allow event probability to be related to patient characteristics, treatment, drug exposure, or disease status. Such outcomes are highly relevant to drug development because they often directly reflect clinical benefit or risk (Holford, 2013) .

In many applications, event risk depends on biomarkers that evolve during treatment. Tumour size, viral load, symptom scores, and other longitudinal measures may reflect changing disease state and prognosis (Liu et al., 2025). The biomarker and TTE processes are interdependent because biomarker deterioration may increase event risk, while occurrence of the event terminates subsequent biomarker observations. This can bias estimation of longitudinal trajectories because high-risk patients contribute shorter follow-up (Kerioui et al., 2022). Joint longitudinal-TTE models address this problem by simultaneously describing biomarker dynamics and event outcomes through shared patient- specific parameters. This differs from sequential approaches, in which the longitudinal model is estimated first and its predictions are subsequently entered into the TTE model. Such approaches may incompletely propagate uncertainty and can introduce bias in longitudinal and survival parameters.

A further challenge in conventional pharmacometric TTE modeling is specification of the hazard function. Parametric models require selection among functions such as exponential, Gompertz, Weibull, log-normal, or log-logistic, each imposing different assumptions about how risk changes over time. Selecting an appropriate hazard can be difficult when empirical hazards are sparse, noisy, or nonmonotonic, while more flexible functions may increase parameter sensitivity and numerical instability (Van Wijk & Simonsson, 2022).

Multi-task logistic regression (MTLR) provides an alternative by directly modeling the survival distribution across ordered time intervals rather than requiring specification of a continuous parametric baseline hazard. Its dependent logistic regressors accommodate censoring and permit predictor effects to vary over time. However, conventional MTLR uses a linear predictor structure (Yu et al., 2011). Neural MTLR improves flexibility by replacing this with a multilayer neural network capable of capturing complex nonlinear relationships. This improved flexibility comes at the cost of reduced interpretability associated with black-box neural networks (Fotso, 2018). The MTLR and its deep learning extensions are used to predict precise, personalized patient survival curves by analyzing complex multi-omics data, clinical registries, and digital pathology images without the rigid constraints of traditional hazard assumptions (Bisaso et al., 2018; Skubleny et al., 2024; Yu et al., 2011).

To address these limitations, we propose replacing the neural network with a dynamic pharmacometric longitudinal model that explicitly represents pharmacodynamics, biomarker trajectories, measurement error, and between-patient variability. To overcome the drawbacks of sequential modeling, the longitudinal and MTLR components are estimated simultaneously, forming a joint longitudinal-MTLR framework with flexible patient-specific survival prediction while avoiding prespecification of a parametric baseline hazard. The present work evaluates this approach first in a controlled simulated dataset and then in a larger controlled phase 3 clinical study dataset involving patients with advanced/metastatic Non-Small Cell Lung Cancer treated with erlotinib.

## Methods

### Study design and data

A two-part methodological evaluation was conducted. The first analysis utilized a simulation- based comparison of conventional joint longitudinal-hazard models, mechanistic MTLR, and hybrid neural-mechanistic MTLR under a common tumour-dynamics model. The second analysis applied the same event-model families to a clinical study dataset to examine estimability and descriptive predictive adequacy under sparse longitudinal sampling.

#### Simulated dataset

The simulated analysis used the synthetic dataset obtained from (Kerioui et al., 2022). . The dataset contained 100 patients and 570 longitudinal SLD observations measured on a nominal 63-day schedule from day 0 to day 693. Individual patients contributed between 1 and 12 SLD observations, with a median of 5, reflecting termination of longitudinal observation following an event. Eighty-one patients experienced an event and 19 were administratively right-censored at day 693. The dataset contained subject identifier, observation time, event or censoring time T, and event indicator δ.

#### Clinical dataset and Progression Free Survival endpoint

The clinical case study used de-identified patient-level data from the control arm of a phase III A6181087/SUN1087 study, NCT00457392, conducted in previously treated advanced non-small- cell lung cancer (Scagliotti et al., 2012). The data obtained from the project data sphere repository (Project Data Sphere, 2015). Progression Free Survival (PFS) was defined from randomization to the first objective disease progression or on-study death. Patients without an event were censored according to the source-study assessment rules. Longitudinal SLD was constructed from target-lesion measurements, and only observations occurring on or before the patient’s PFS event or censoring time were retained. A usable baseline SLD measurement was required for inclusion.

The final clinical modeling cohort contained 453 patients, of whom 366 experienced a PFS event and 87 were censored. A total of 1,346 SLD observations were available, including 21 exact zero SLD values. Sampling was sparse, with 98 patients contributing baseline SLD only and 286 of 453 patients contributing two or fewer SLD observations.

### Longitudinal model

For the simulated dataset, tumour dynamics were described using the nonlinear shrinkage–regrowth model in which an initially treatment-sensitive component declines while a resistant component subsequently grows.

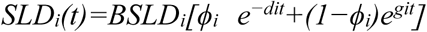

where BSLD_i_ is baseline tumour size, ϕ_i_ the treatment-sensitive fraction, d_i_ the shrinkage rate, and g_i_ the regrowth rate. This formulation represents initial tumour reduction followed by regrowth ^(^Liu et al., 2025^)^. A proportional residual-error model was used for observed SLD. A nonlinear mixed-effects model estimated population parameters, interindividual variability, and residual error. The same longitudinal model was used for all event models. The same longitudinal model was used for all retained event models to ensure that comparisons reflected differences in the event-model formulation rather than differences in the representation of tumour dynamics.

For the clinical dataset, the same parent equation was retained but the sparse pre-PFS longitudinal data did not support separate estimation of d, g, and ϕ with patient-specific variability. The primary clinical analysis therefore used the nested shrinkage-plus-plateau form obtained by fixing g = 0:

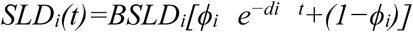

Only baseline tumour burden was assigned interindividual variability, log(BSLD_i_) = log(BSLD) + η_i_, with η_i_ distributed N(0, ω²). The population parameters d and ϕ were estimated once in a dedicated longitudinal-only model and then fixed during event-model comparison. Because the clinical dataset contained zero SLD values and scale-dependent variability, the observation model was fitted on the log1p scale:

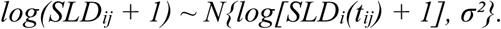

This transformation retained the structural SLD trajectory on the original millimeter scale while stabilizing the residual distribution and allowing exact zero SLD observations.

### Multi-Task Logistic Regression

Multi-Task Logistic Regression is a discrete-time survival-distribution model consisting of a sequence of dependent local logistic regressors (Yu et al., 2011). Unlike conventional parametric TTE models, MTLR directly models the survival distribution over ordered time intervals rather than specifying a parametric baseline hazard. Follow-up was divided into intervals

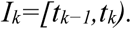

using ordered cut points

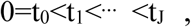

For conventional MTLR, the local output at time interval _j_ is

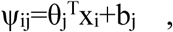

where x_i_ represents patient predictors and θ_j_ and b_j_ are interval-specific coefficients and intercepts. The cumulative interval score

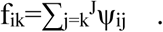

These scores are transformed into a valid discrete event-time distribution using

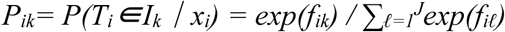

Consequently,

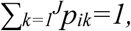

and survival probabilities were obtained as tail sums,

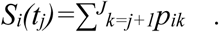

This construction allows time-varying covariate effects while producing valid monotone survival distributions.

For right-censored observations, likelihood contributions were obtained by summing probabilities across all compatible future event intervals. Thus, if patient i was known to remain event-free until censoring time c_i_ ,

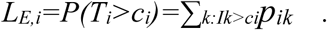

Neural-MTLR replaces the linear predictor with a multilayer neural network,

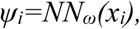

while retaining the original MTLR probability and censoring structure (Fotso, 2018). This enables nonlinear predictor-survival relationships but reduces mechanistic interpretability.

### Proposed mechanistic MTLR

To retain the survival-distribution advantages of MTLR while improving biological interpretability, we proposed replacement of the neural representation with the model-predicted latent tumour trajectory. For the simulated analysis, at MTLR time point t_j_ ,

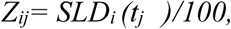

For the clinical analysis, the mechanistic association was defined on the centered log-transformed SLD scale:

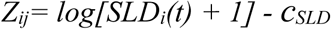

where c_SLD was the median baseline log(SLD + 1) (Zhudenkov et al., 2022). Clinical MTLR cut points were 28, 56, 84, 112, 168, 224, 336, 504, and 720 days and

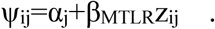

where αj represents an interval-specific baseline score and β_MTLR_ quantifies the relationship between the current latent tumour burden and the event-time distribution. Scaling SLD by 100 was used to improve parameter scaling and numerical stability. These scores were converted to event-interval probabilities using the standard MTLR normalization above.

Unlike a sequential analysis, the patient-specific random effects generating SLDi (t) simultaneously entered the longitudinal and event components. The joint likelihood was

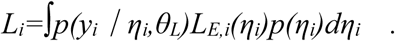

Thus, survival information contributed to estimation of the shared latent disease trajectory. This distinguishes joint estimation from two-stage approaches, which may introduce bias when the longitudinal and survival processes are associated.

### Hybrid neural–mechanistic MTLR

The hybrid model retained the mechanistic relationship while adding a neural component to capture additional nonlinear information from the observed longitudinal measurements.

Observed SLD was aligned to the MTLR time grid using last-observation-carried-forward and standardized using the mean and standard deviation estimated from the training data. For each profiled neural basis parameter W1 , with candidate values 0.35, 0.60, 1.00, and 1.50, the transformed input *h_ij_=tanh(W1z_ij_)* was orthogonalized with respect to the intercept and linear observed-SLD effect by subtracting its fitted linear projection. The resulting residual basis was normalized to unit root-mean-square magnitude, yielding q_ij_, so that the neural contribution represented nonlinear information not explained by a linear SLD effect.

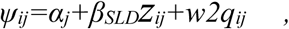

w2 quantified the additional nonlinear observed-SLD contribution.

In the clinical analysis, the hybrid extension used the same principle on observed log(SLD + 1) history with W = 1.0. The nonlinear basis was centered, orthogonalized against its linear component, bounded, and normalized. The hybrid clinical fit was conditional on the population longitudinal parameters estimated by mechanistic MTLR, while the subject-specific BSLD random effect remained integrated by Laplace approximation.

### Conventional joint hazard comparator

For comparison, the conventional joint model linked the same tumour trajectory but alternative parametric baseline hazard functions. The candidate hazard structures comprised the exponential, Gompertz, Weibull, log-normal, log-logistic, and periodic/circadian functions commonly considered in pharmacometric time-to-event analyses [<u>1</u>]. The latent SLD trajectory was linked to a continuous-time hazard according to

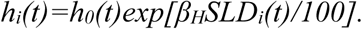

where h_i_(t) is the individual hazard at time t, h_0_(t) is the model-specific baseline hazard, and *β_H_* quantifies the association between the current mechanistically predicted SLD and event risk. Thus, the longitudinal submodel and longitudinal–event association structure were held constant, while only the functional form of the baseline hazard was varied.

The baseline hazards were specified as exponential,

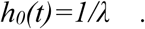

Gompertz,

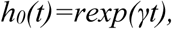

Weibull,

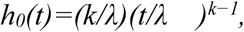

log-normal,

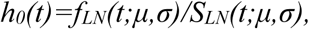

and log-logistic,

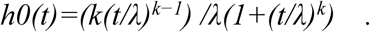

The periodic model was implemented as

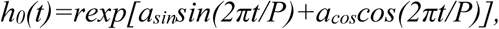

using sine and cosine coefficients to avoid direct estimation of a bounded phase parameter. Thus, within each dataset the longitudinal structure and longitudinal-event association were held constant while the functional form of the baseline hazard was varied. The corresponding survival function was

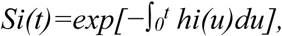

and the event contribution to the likelihood was

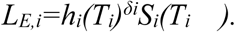

### Estimation, diagnostics and validation

The longitudinal model was first fitted in R with the SAEMIX package to estimate population parameters, variability, residual error, and empirical Bayes trajectories (Comets et al., 2026). These estimates provided starting values for joint estimation. The joint hazard, mechanistic MTLR, and hybrid models were then estimated in R with Template Model Builder (TMB) using automatic differentiation and Laplace approximation over random effects (Kristensen et al., 2016). Convergence required successful optimizer termination, finite objective function and gradients, plausible parameter estimates, and a numerically valid Hessian. Models failing these criteria were not accepted for predictive comparison. Model evaluation included observed-versus-predicted SLD, residuals versus time and predictions and longitudinal visual predictive checks.

For the survival component, the visual predictive check compared model-predicted probabilities of remaining event-free with the empirical survival distribution from a Kaplan-Meier curve. A true model-specific VPC was generated using 300 replicate datasets per converged clinical model. For each continuous-time hazard model, a new BSLD random effect was simulated for each patient, the latent SLD trajectory and individual hazard were reconstructed, and an event time was generated by inversion of the cumulative hazard. Independent censoring times were sampled from the empirical censoring distribution estimated by reverse Kaplan-Meier. For mechanistic and hybrid MTLR, event intervals were sampled from the fitted MTLR interval probabilities; a uniform time within the selected interval was used only for visualization. The survival VPC ribbon therefore represents the 2.5th to 97.5th percentiles of model-simulated Kaplan-Meier curves rather than a confidence interval around the observed Kaplan-Meier estimator.

For the simulated dataset, predictive performance was assessed using five-fold patient-level cross- validation with identical folds for all models. For each held-out patient, latent states were estimated using longitudinal SLD history only; event time and event status were excluded to prevent leakage. Calibration and discrimination were evaluated independently because a model may rank high-risk patients correctly while providing poorly calibrated survival probabilities. Dynamic prediction performance was therefore assessed using time-dependent discrimination and probability-scoring metrics. Dynamic predictions were evaluated over 0–189, 189–378, and 378–567 days using a landmark framework. At landmark time s, only longitudinal observations available up to s,

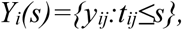

were available for patient-specific latent-state estimation. The resulting individual trajectory was propagated beyond the landmark to obtain future SLD values and event probabilities.

For the hazard model, conditional survival was calculated as

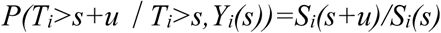

For the MTLR models, interval probabilities over the future prediction window were obtained from the MTLR distribution and corresponding survival probabilities were calculated by tail summation. Thus, additional longitudinal observations could dynamically modify the predicted future event distribution, consistent with the principle of dynamic joint-model prediction. Predictive discrimination was quantified using the dynamic inverse-probability-of-censoring-weighted (IPCW) AUC (Blanche et al., 2015; Dong et al., 2020; Graf et al., 1999; Heagerty et al., 2000; Yang et al., 2025). Probability accuracy was assessed using the dynamic IPCW Brier score and integrated Brier score (IBS) (Brier, 1950; Dong et al., 2020; Prince et al., 2025), while a common interval-based negative log score assessed the probability assigned to the observed event interval (Bracher et al., 2021). Calibration was examined by comparing predicted and observed survival probabilities. Higher AUC indicated better discrimination, whereas lower Brier score, IBS, and negative log score indicated better predictive performance. The clinical case study was was not subjected to the five-fold dynamic cross-validation used for the simulated dataset.

## Results

### Simulated-data analysis

All eight joint models converged satisfactorily and produced similar longitudinal parameter estimates despite substantial differences in their event-model formulations (Table 1). Baseline SLD estimates ranged from 116.888 to 116.938 and residual-error estimates from 0.1007 to 0.1011. Estimates of the shrinkage rate, regrowth rate, and responsive fraction were also similar across models, particularly among the six parametric hazard formulations. The mechanistic and hybrid MTLR models produced slightly higher shrinkage and regrowth rate estimates but retained comparable baseline SLD, responsive fraction, and residual error.

**Table 1.** Development-stage event-model parameter estimates.

| Model | BSLD | Shrinkage rate, d | Regrowth rate, g | Responsive fraction, $\phi$ | Residual error, $\sigma$ | Current-SLD association, βSLD | Additional event parameter(s) |
| --- | --- | --- | --- | --- | --- | --- | --- |
| Exponential joint hazard | 116.930 | 0.003785 | 0.003084 | 0.6628 | 0.1010 | 0.988 | — |
| Gompertz joint hazard | 116.919 | 0.003784 | 0.003085 | 0.6631 | 0.1010 | 0.986 | Rate = 0.000633;<br>$\gamma/100$ days = -0.0309 |
| Weibull joint hazard | 116.923 | 0.003785 | 0.003085 | 0.6629 | 0.1010 | 0.988 | Scale = 1744.58;<br>shape = 0.984 |
| Log-normal joint hazard | 116.938 | 0.003784 | 0.003085 | 0.6633 | 0.1011 | 0.993 | $\mu = 7.501$; $\sigma_{LN} = 1.900$ |
| Log-logistic joint hazard | 116.933 | 0.003784 | 0.003083 | 0.6630 | 0.1010 | 0.982 | Scale = 1351.26;<br>shape = 1.063 |
| Circadian joint hazard | 116.888 | 0.003786 | 0.003091 | 0.6634 | 0.1010 | 0.999 | Rate = 0.000532;<br>amplitude = 0.490; period = 141.1 days |
| Mechanistic MTLR | 116.893 | 0.003958 | 0.003772 | 0.6724 | 0.1007 | 0.388 | — |
| Hybrid neural-mechanistic MTLR | 116.929 | 0.003956 | 0.003745 | 0.6678 | 0.1008 | 0.502 | Neural residual weight, $w_2 = 0.655$ |

Note. BSLD, d, g, $\phi$ , and $\sigma$ are the estimated longitudinal population parameters. Interindividual variability parameters were fixed to the values obtained from the longitudinal SAEMIX fit and therefore were not re-estimated separately for each joint model. The $\beta$ SLD estimates from hazard and MTLR formulations operate on different event-model scales and should not be compared directly by magnitude.

The estimated SLD association was positive in all event models. The hazard-based models gave similar coefficients, ranging from 0.982 to 0.999. The MTLR coefficients are on a different event- model scale and should therefore not be interpreted as directly comparable in magnitude with the hazard-model coefficients. Among the five additional parametric hazard models, the circadian model had the lowest AIC (6179.2), whereas the log-logistic model had the lowest subject-level BIC (6204.4).

Longitudinal goodness-of-fit was nearly identical across the eight models (Table 2). RMSE ranged from 10.084 to 10.165 and MAE from 6.900 to 6.930. Observed SLD values closely followed individual predictions for all models, with modest deviation only at the largest SLD values (Figure 1A–H). Weighted residuals were generally centered around zero, with similar minor temporal trends across models (Figure 1I–P).

**Figure 1.**
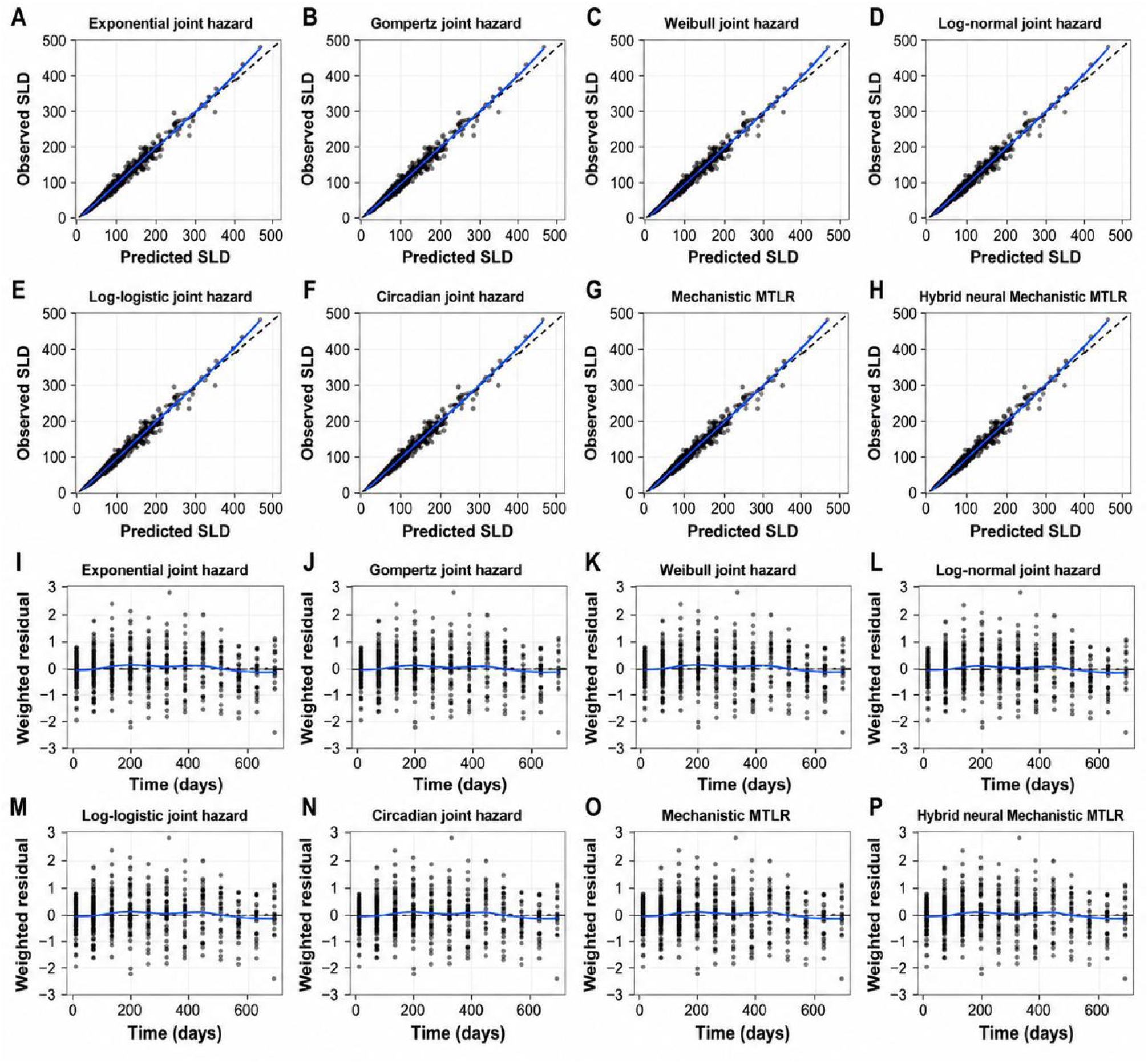
Longitudinal goodness-of-fit diagnostics for the joint time-to-event models. Panels A–H show observed versus individual predicted sum of longest diameters (SLD) for the exponential, Gompertz, Weibull, log-normal, log-logistic, circadian, mechanistic MTLR, and hybrid neural–mechanistic MTLR models, respectively. The dashed diagonal line represents the line of identity and the blue line represents the LOESS trend. Panels I–P show weighted residuals versus time for the corresponding models. The dashed horizontal line indicates zero residual and the blue LOESS curve summarizes temporal trends in residuals.

**Table 2.** Development-stage longitudinal goodness-of-fit and event calibration.

| Model | SLD RMSE | SLD MAE | Mean absolute calibration error | Event VPC coverage |
| --- | --- | --- | --- | --- |
| Exponential joint hazard | 10.102 | 6.900 | 0.0280 | 8/8 |
| Gompertz joint hazard | 10.115 | 6.905 | 0.0222 | 8/8 |
| Weibull joint hazard | 10.105 | 6.902 | 0.0259 | 8/8 |
| Log-normal joint hazard | 10.165 | 6.916 | 0.0151 | 8/8 |
| Log-logistic joint hazard | 10.114 | 6.903 | 0.0227 | 8/8 |
| Circadian joint hazard | 10.130 | 6.907 | 0.0206 | 8/8 |
| Mechanistic MTLR | 10.084 | 6.908 | 0.0167 | 8/8 |
| Hybrid neural-mechanistic MTLR | 10.145 | 6.930 | 0.0182 | 8/8 |

Visual predictive checks further supported adequate model performance (Figure 2). For all eight models, the observed 10th, 50th, and 90th percentiles of SLD were contained within their corresponding 95% simulation intervals at all 12 evaluation times (Figure 2A). Similarly, the observed Kaplan–Meier event-free probabilities were contained within the 95% simulation intervals at all eight evaluation times for every model (Figure 2B).

**Figure 2.**
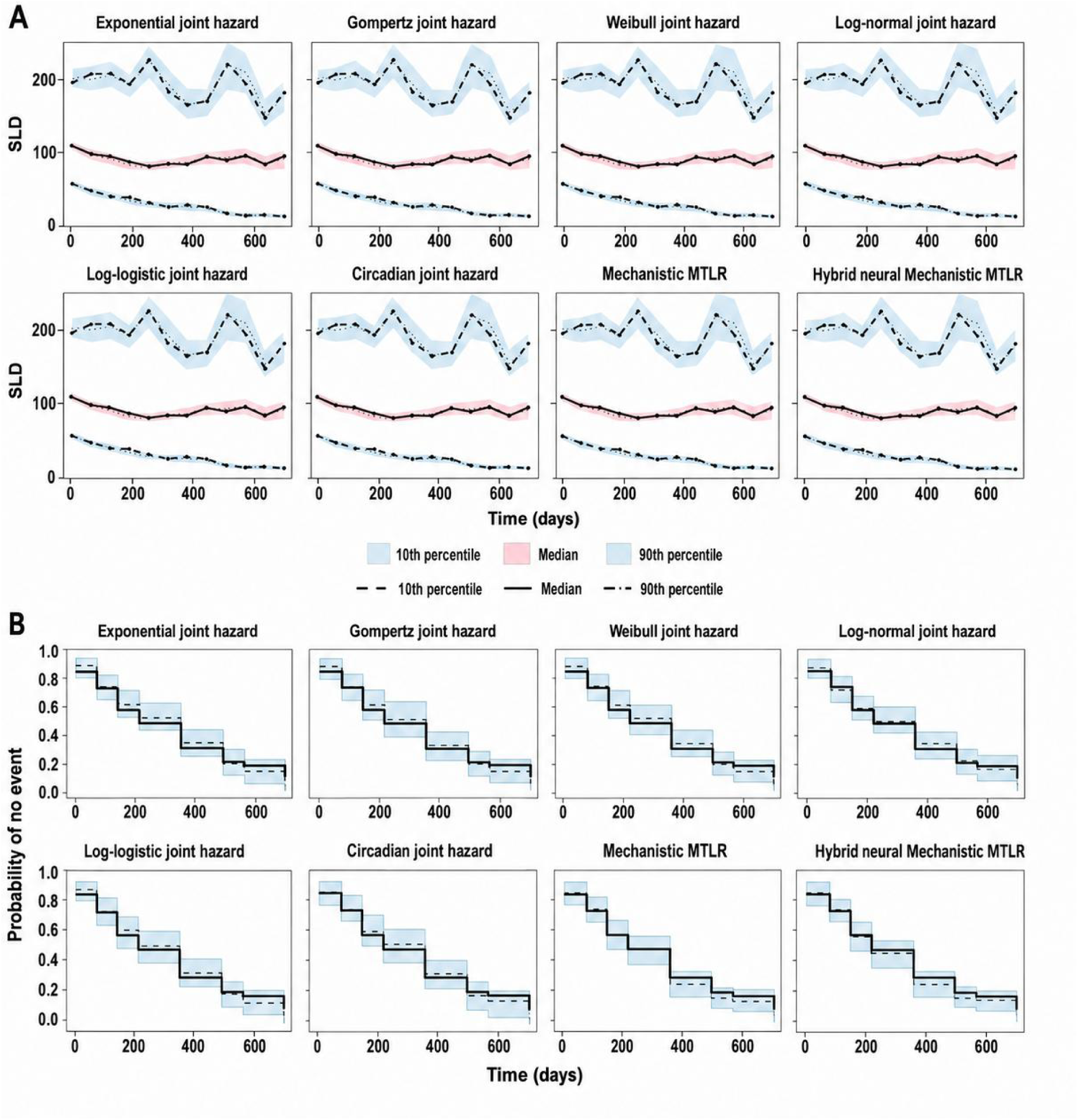
Visual predictive checks for the longitudinal and event components of the joint models. (A) Longitudinal VPCs of SLD over time for the exponential, Gompertz, Weibull, log-normal, log-logistic, circadian, mechanistic MTLR, and hybrid neural–mechanistic MTLR models. Shaded regions represent the 95% simulation intervals for the 10th and 90th percentiles in light blue and the median in light pink, with observed and simulated quantiles shown by the corresponding lines. (B) Event VPCs showing the probability of remaining event-free over time. Solid step lines represent the observed Kaplan–Meier estimates, dashed step lines represent the median simulated probabilities, and light- blue stepped bands represent the 95% simulation intervals.

Five-fold cross-validation demonstrated a clear distinction between discrimination and probabilistic prediction across the event-model formulations (Table 3; Figure 3). During the 0–189-day window, all six conventional hazard models and mechanistic MTLR produced the same AUC of 0.676, whereas the hybrid model had a lower AUC of 0.573. The log-normal hazard provided the lowest Brier score in this interval (0.2177).

**Figure 3.**
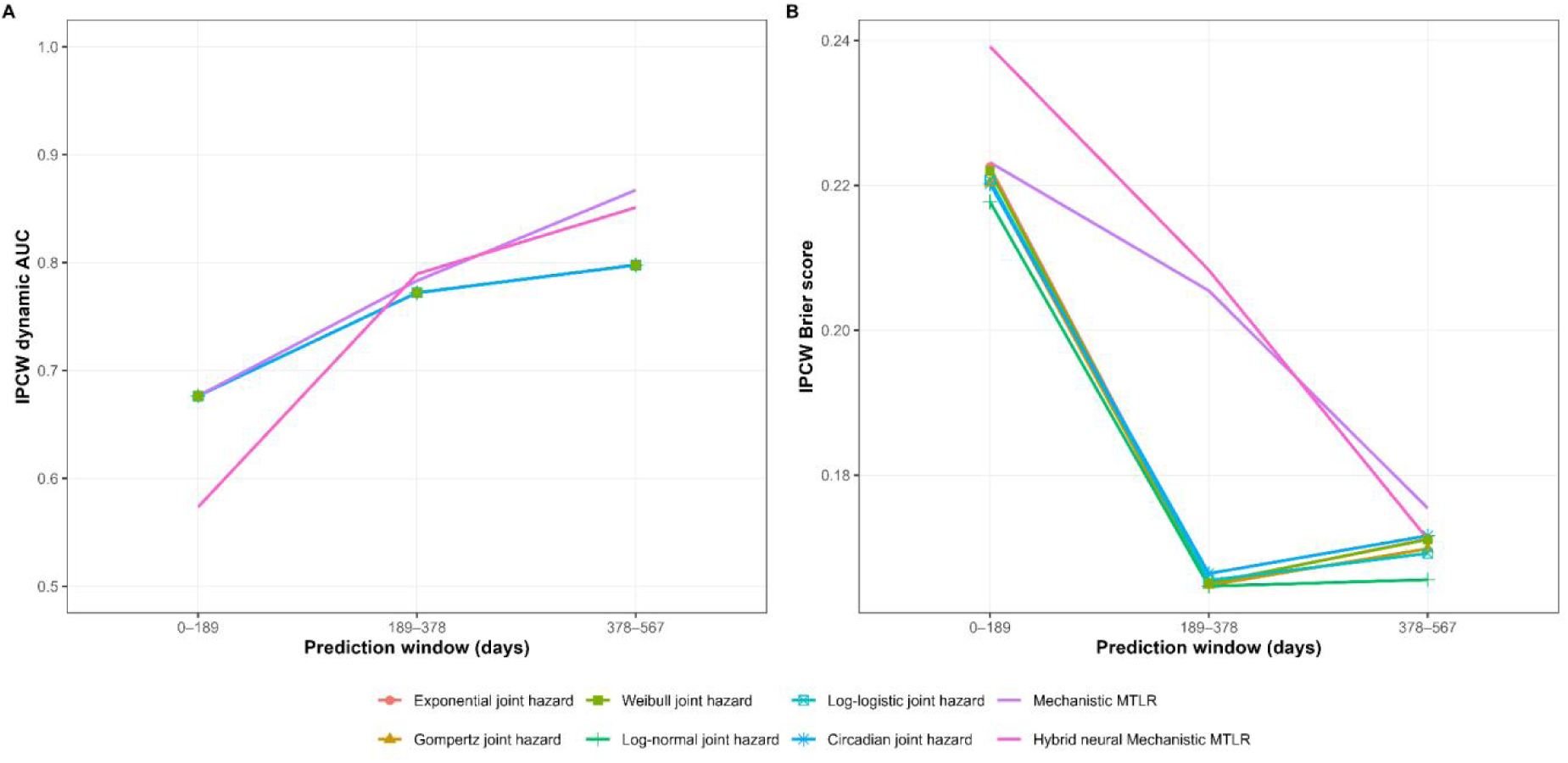
Five-fold cross-validated dynamic predictive performance of the joint time-to-event models. (A) Inverse probability of censoring weighted (IPCW) dynamic area under the receiver operating characteristic curve (AUC) and (B) IPCW Brier score across the 0–189, 189–378, and 378–567-day prediction windows for the six conventional joint hazard models, mechanistic MTLR, and hybrid neural–mechanistic MTLR. Higher AUC indicates better discrimination, whereas lower Brier scores indicate better probabilistic predictive performance.

**Table 3.** Five-fold cross-validated dynamic predictive performance.

| Window, days | Model | AUC | Brier score | Predicted survival | Observed survival |
| --- | --- | --- | --- | --- | --- |
| 0-189 | Exponential joint hazard | 0.676 | 0.2225 | 0.679 | 0.590 |
| 0-189 | Gompertz joint hazard | 0.676 | 0.2203 | 0.669 | 0.590 |
| 0-189 | Weibull joint hazard | 0.676 | 0.2220 | 0.676 | 0.590 |
| 0-189 | Log-normal joint hazard | 0.676 | 0.2177 | 0.649 | 0.590 |
| 0-189 | Log-logistic joint hazard | 0.676 | 0.2208 | 0.669 | 0.590 |
| 0-189 | Circadian joint hazard | 0.676 | 0.2202 | 0.666 | 0.590 |
| 0-189 | Mechanistic MTLR | 0.676 | 0.2232 | 0.670 | 0.590 |
| 0-189 | Hybrid neural-mechanistic MTLR | 0.573 | 0.2391 | 0.649 | 0.590 |
| 189-378 | Exponential joint hazard | 0.772 | 0.1651 | 0.596 | 0.610 |
| 189-378 | Gompertz joint hazard | 0.772 | 0.1648 | 0.598 | 0.610 |
| 189-378 | Weibull joint hazard | 0.772 | 0.1651 | 0.598 | 0.610 |
| 189-378 | Log-normal joint hazard | 0.772 | 0.1647 | 0.607 | 0.610 |
| 189-378 | Log-logistic joint hazard | 0.772 | 0.1655 | 0.594 | 0.610 |
| 189-378 | Circadian joint hazard | 0.772 | 0.1664 | 0.594 | 0.610 |
| 189-378 | Mechanistic MTLR | 0.783 | 0.2055 | 0.511 | 0.610 |
| 189-378 | Hybrid neural-mechanistic MTLR | 0.789 | 0.2083 | 0.484 | 0.610 |
| 378-567 | Exponential joint hazard | 0.798 | 0.1712 | 0.645 | 0.722 |
| 378-567 | Gompertz joint hazard | 0.798 | 0.1698 | 0.662 | 0.722 |
| 378-567 | Weibull joint hazard | 0.798 | 0.1711 | 0.649 | 0.722 |
| 378-567 | Log-normal joint hazard | 0.798 | 0.1656 | 0.703 | 0.722 |
| 378-567 | Log-logistic joint hazard | 0.798 | 0.1692 | 0.661 | 0.722 |
| 378-567 | Circadian joint hazard | 0.798 | 0.1717 | 0.679 | 0.722 |
| 378-567 | Mechanistic MTLR | 0.867 | 0.1754 | 0.726 | 0.722 |
| 378-567 | Hybrid neural-mechanistic MTLR | 0.851 | 0.1712 | 0.727 | 0.722 |

**Figure 4.**
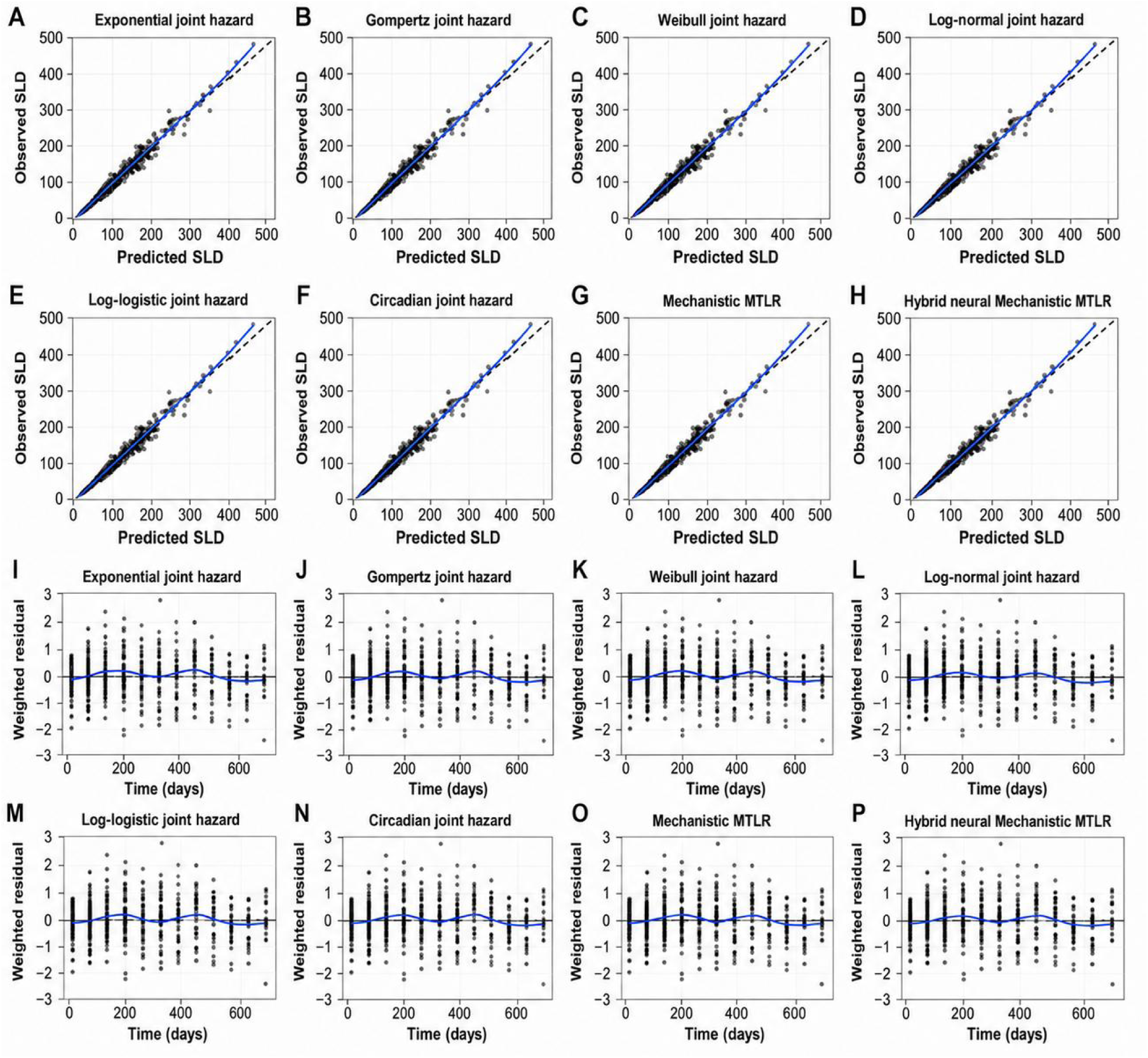
Longitudinal goodness-of-fit diagnostics for the clinical PFS case study. Panels A-H show observed versus individual predicted SLD for the exponential, Gompertz, Weibull, log-normal, log-logistic, periodic, mechanistic MTLR, and hybrid neural-mechanistic MTLR models. The dashed diagonal line is the line of identity and the blue line is a locally smoothed trend. Panels I-P show standardized residuals versus time for the corresponding models; the horizontal reference line denotes zero residual.

**Figure 5.**
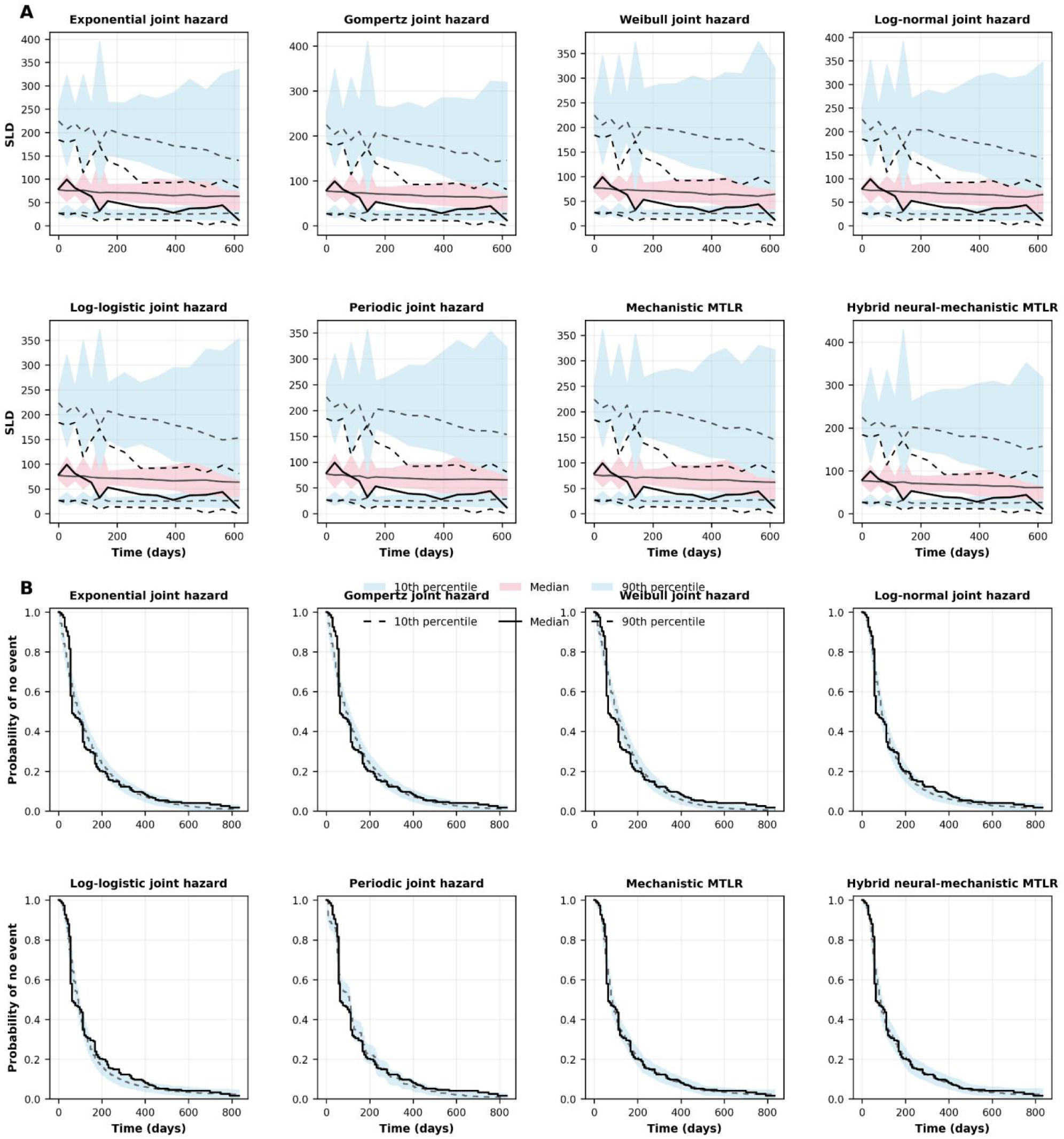
Visual predictive checks for the clinical PFS case study. (A) Longitudinal SLD VPCs. Shaded regions show 95% simulation intervals for the 10th and 90th percentiles in light blue and the median in light pink; observed and simulated quantiles are shown by the corresponding lines. (B) True model-specific PFS VPCs. The black step line is the observed Kaplan-Meier curve, the dashed line is the median simulated Kaplan-Meier curve, and the light- blue band is the 95% interval across 300 PFS datasets simulated from the model shown in each panel.

At 189–378 days, all conventional hazard functions again produced the same AUC of 0.772, whereas discrimination was higher for mechanistic MTLR (0.783) and highest for the hybrid model (0.789). In contrast, the conventional hazard models had lower Brier scores, with the log-normal model providing the lowest value (0.1647). At 378–567 days, the six hazard models again had identical discrimination (AUC 0.798), while the mechanistic MTLR achieved the highest AUC of 0.867 and the hybrid model an AUC of 0.851. The log-normal model again produced the lowest Brier score (0.1656).

Across the complete 0–567-day evaluation period, differences in IBS among the six conventional hazard formulations were small, ranging from 0.1928 to 0.1943 (Table 4). The log-normal joint hazard model had the lowest overall IBS (0.1928), followed by log-logistic (0.1933) and Gompertz (0.1935). Mechanistic MTLR had a somewhat higher IBS of 0.2011, whereas the hybrid model had the highest value of 0.2091. A different pattern was observed for event-interval prediction. Mechanistic MTLR achieved the lowest mean negative log score (2.362), followed by the hybrid model (2.538).

**Table 4.**
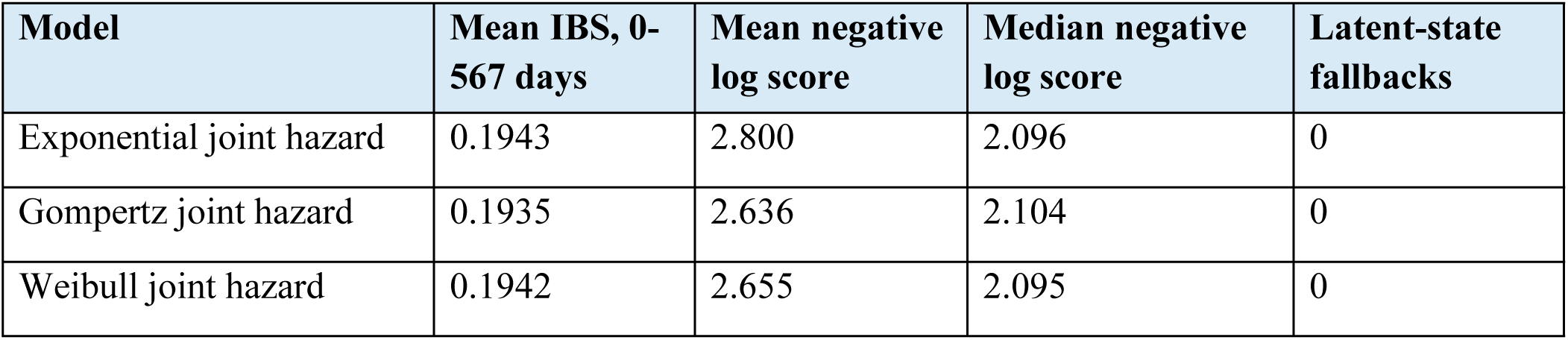
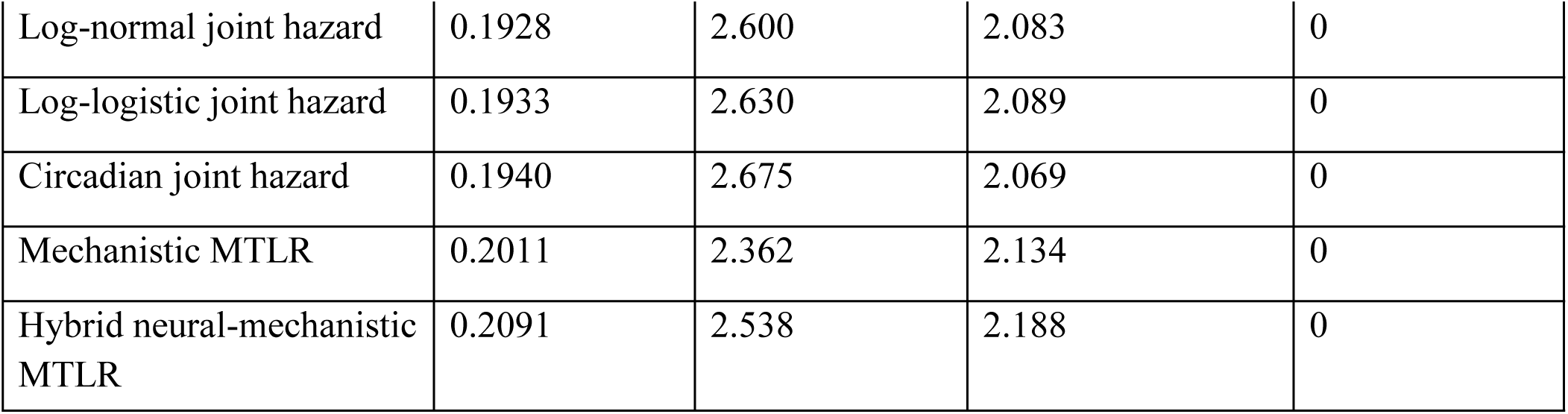
Overall cross-validated predictive performance.

| Model | Mean IBS, 0-567 days | Mean negative log score | Median negative log score | Latent-state fallbacks |
| --- | --- | --- | --- | --- |
| Exponential joint hazard | 0.1943 | 2.800 | 2.096 | 0 |
| Gompertz joint hazard | 0.1935 | 2.636 | 2.104 | 0 |
| Weibull joint hazard | 0.1942 | 2.655 | 2.095 | 0 |
| Log-normal joint hazard | 0.1928 | 2.600 | 2.083 | 0 |
| Log-logistic joint hazard | 0.1933 | 2.630 | 2.089 | 0 |
| Circadian joint hazard | 0.1940 | 2.675 | 2.069 | 0 |
| Mechanistic MTLR | 0.2011 | 2.362 | 2.134 | 0 |
| Hybrid neural-mechanistic MTLR | 0.2091 | 2.538 | 2.188 | 0 |

**Table 5.**
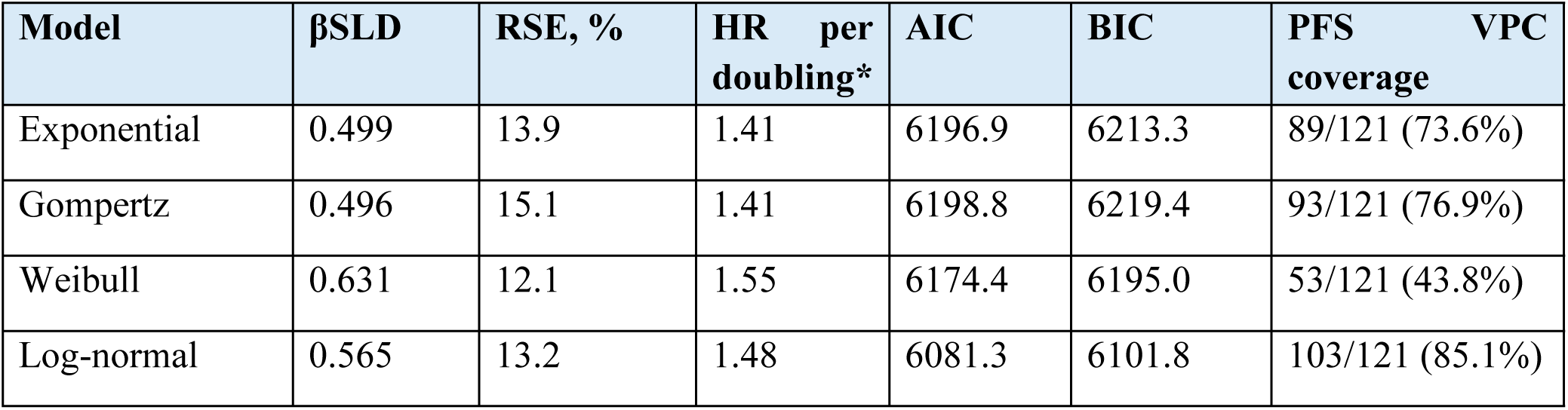
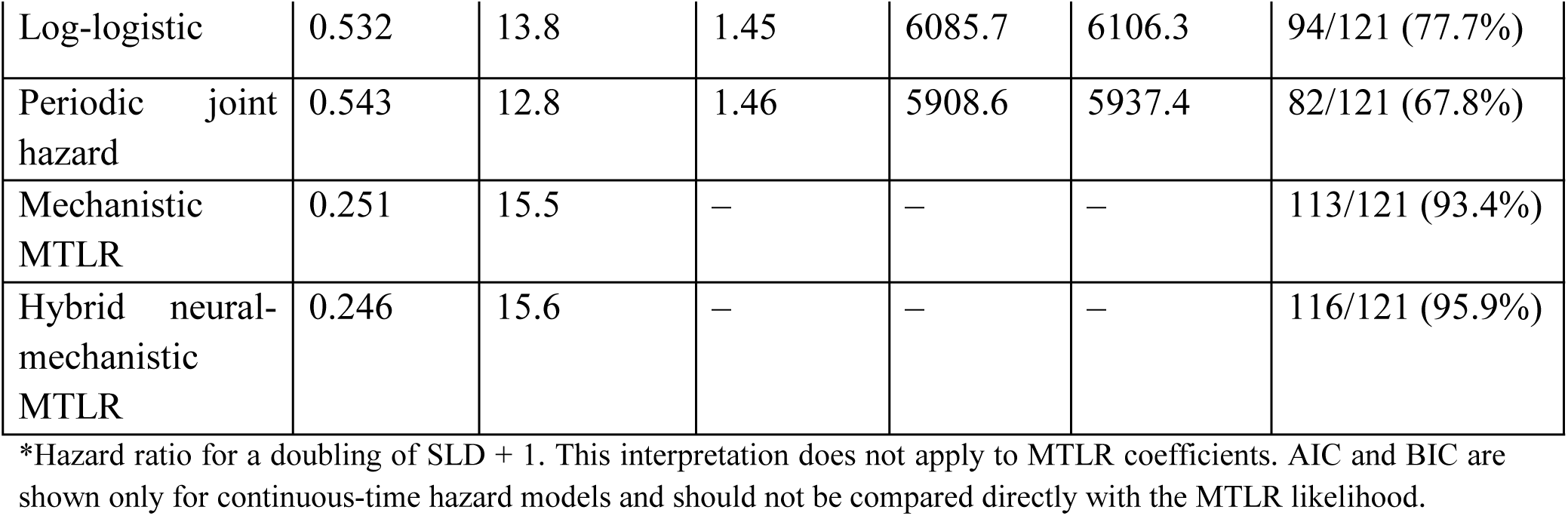
Clinical-data development-stage event-model results.

**Table 6.** Clinical-data diagnostic summary.

| Model | SLD RMSE | SLD MAE | KM MAE | PFS VPC coverage |
| --- | --- | --- | --- | --- |
| Exponential | 0.2550 | 0.1491 | 0.0264 | 89/121 (73.6%) |
| Gompertz | 0.2550 | 0.1491 | 0.0255 | 93/121 (76.9%) |
| Weibull | 0.2553 | 0.1496 | 0.0341 | 53/121 (43.8%) |
| Log-normal | 0.2552 | 0.1491 | 0.0201 | 103/121 (85.1%) |
| Log-logistic | 0.2551 | 0.1490 | 0.0215 | 94/121 (77.7%) |
| Periodic joint hazard | 0.2551 | 0.1491 | 0.0238 | 82/121 (67.8%) |
| Mechanistic MTLR | 0.2554 | 0.1490 | 0.0117 | 113/121 (93.4%) |
| Hybrid neural-mechanistic MTLR | 0.2554 | 0.1490 | 0.0121 | 116/121 (95.9%) |

SLD RMSE and MAE are calculated on the $\log(\text{SLD} + 1)$ observation scale. KM MAE is the mean absolute difference between the median simulated Kaplan-Meier curve and the observed Kaplan-Meier curve over the VPC evaluation grid. PFS VPC coverage is a development-stage diagnostic and is not external validation.

Overall, changing the parametric baseline hazard had little effect on discrimination, as all six conventional hazard formulations produced identical AUCs within each prediction window. Hazard-function choice had a greater effect on calibration and probabilistic accuracy, with the log-normal model consistently producing the lowest dynamic Brier scores and the lowest overall IBS. In contrast, mechanistic MTLR provided superior event-interval probability prediction and greater discrimination at the later prediction landmarks. The hybrid neural–mechanistic model improved discrimination relative to the conventional hazard models after the baseline prediction window but did not consistently improve probabilistic accuracy relative to mechanistic MTLR.

### Clinical-data analysis

All eight clinical models met the prespecified numerical criteria, with optimizer convergence code 0, maximum absolute gradients between 0.0047 and 0.0437, and positive-definite Hessians. The dedicated longitudinal fit estimated d = 0.002027 day^-1^ and ϕ = 0.2544, while g was fixed at 0. The BSLD random-effect standard deviation was ω = 0.7762. These longitudinal shape parameters were then fixed during the event-model comparison.

The fitted population baseline SLD was highly consistent across the seven fully joint clinical event models, ranging from 76.79 to 77.01 mm, and the residual standard deviation on the log(SLD + 1) scale ranged from 0.3067 to 0.3069. Conditional longitudinal goodness-of-fit was also nearly identical across models, with RMSE 0.2550-0.2554, MAE 0.1490-0.1496, mean residual approximately -0.002, and observed-versus-predicted correlations of approximately 0.964. The SLD-event association was positive in every clinical model. Among the continuous-time hazard models, β_SLD_ ranged from 0.496 to 0.631 with RSEs of 12.1%-15.1%. Because the clinical hazard predictor was centered log(SLD + 1), these coefficients corresponded to an approximate 1.41- to 1.55-fold increase in instantaneous PFS event hazard for a doubling of SLD + 1. The mechanistic and hybrid MTLR coefficients were 0.251 and 0.246, respectively, and should not be interpreted as hazard ratios. The hybrid nonlinear weight was small, w_SLD_ = 0.0291, SE 0.0241, RSE 82.9%.

Within the continuous-time clinical hazard models, the periodic formulation had the lowest AIC, 5908.6, followed by log-normal, 6081.3, and log-logistic, 6085.7. The periodic model estimated a period of approximately 57.75 days and a sine-cosine amplitude of approximately 1.52, corresponding to a large peak-to-trough baseline hazard contrast. Given the scheduled nature of radiographic PFS assessment, this periodicity is more plausibly interpreted as an assessment or ascertainment effect than as a biological rhythm. Among smooth non-periodic hazard families, the log-normal model had the lowest AIC.

The clinical longitudinal VPC revealed a more important limitation than was apparent from individual goodness-of-fit. Across the 15 evaluable time windows, the observed 10th percentile was contained within its 95% simulation interval at only 4/15 windows for all models; median coverage ranged from 5/15 to 7/15, and 90th-percentile coverage was 4/15 for most models and 5/15 for the exponential model. Thus, the identifiable one-random-effect longitudinal model described individual predictions adequately but did not fully reproduce population-level longitudinal heterogeneity.

The model-specific clinical PFS VPCs showed stronger descriptive calibration for the MTLR formulations than for most parametric hazards. The observed Kaplan-Meier curve was contained within the simulated 95% interval at 113/121 evaluation times, 93.4%, for mechanistic MTLR and 116/121, 95.9%, for the hybrid model. Corresponding coverage was 103/121, 85.1%, for log- normal; 94/121, 77.7%, for log-logistic; 93/121, 76.9%, for Gompertz; 89/121, 73.6%, for exponential; 82/121, 67.8%, for periodic; and 53/121, 43.8%, for Weibull. Mechanistic MTLR also had the smallest mean absolute difference between the median simulated and observed Kaplan-Meier curves, 0.0117, closely followed by hybrid MTLR, 0.0121.

## Discussion

Joint models provide a well-established framework for linking an underlying longitudinal biomarker trajectory with an event process through shared individual-level information, including in nonlinear pharmacometric settings (Tsiatis & Davidian, 2004; Van Wijk & Simonsson, 2022; Zhudenkov et al., 2022). This study evaluated whether mechanistic MTLR can provide a practical alternative to conventional hazard-based time-to-event modeling when integrated with a mechanistic longitudinal model. The simulated dataset provided a controlled comparison in which the longitudinal mechanism was shared across event models, while the added clinical case study tested whether the same modeling concepts could be estimated and diagnostically evaluated in a substantially larger patient-level clinical PFS dataset with sparse and irregular longitudinal SLD sampling. Taken together, the two analyses support MTLR as a complementary event-model formulation rather than a universally superior replacement for parametric hazards.

In the simulated-data setting, changing the baseline hazard family had little effect on discrimination because all six conventional hazard models produced identical dynamic AUC values within each landmark window. Hazard-family choice nevertheless affected probabilistic accuracy, with the log-normal model producing the lowest dynamic Brier scores and the lowest overall IBS. Mechanistic MTLR showed a different strength. It achieved the lowest event-interval negative log score and superior discrimination at the latest prediction window, where AUC was 0.867 compared with 0.798 for each conventional hazard model. This separation reinforces that discrimination, probability accuracy, and event-interval probability assignment measure different aspects of predictive performance (Steyerberg et al., 2010).

The clinical analysis provided complementary evidence under a more difficult longitudinal sampling pattern. All eight models converged after the longitudinal component was reparameterized to match the information content of the clinical data. The full shrinkage-regrowth formulation used in the simulated experiment could not be supported with patient-specific d, g, and ϕ when most clinical patients had two or fewer pre-PFS SLD measurements. Fixing g at zero, estimating d and ϕ once longitudinally, and retaining only a BSLD random effect produced stable estimation across all event models. This illustrates an important pharmacometric principle. A mechanistic structural equation may remain scientifically useful while its random-effects structure must be adapted to the identifiability supported by the observed design (Duffull et al., 2025).

Conventional parametric TTE modeling requires selection and parameterization of a baseline hazard function. This choice can be consequential because different hazard functions impose different assumptions regarding how event risk changes with time, while more flexible functions may introduce additional estimation and model-selection complexity (Van Wijk & Simonsson, 2022). The present comparison illustrates this issue directly. In the simulated data analysis, although all six hazard models adequately described the data, their probabilistic predictive performance differed. The log-normal model consistently produced the lowest dynamic Brier scores and the lowest overall integrated Brier score, whereas the exponential model was somewhat less competitive. Thus, selecting an appropriate baseline hazard function can improve prediction even when the same longitudinal trajectory and biomarker-event relationship are retained. The clinical results reinforced this concern motivating the comparison and choice of parametric event-model formulations. The periodic hazard had the lowest AIC among the continuous-time clinical hazards, but it estimated a period of approximately 58 days with a very large peak-to-trough hazard contrast. In a PFS dataset based on scheduled radiographic assessments, that pattern is more plausibly related to the timing of tumour assessment and progression ascertainment than to a biological periodic process. Flexible hazard functions can therefore absorb features of the observation process as well as the underlying event process. For this reason, the periodic model should be interpreted as an assessment-process sensitivity model. Among smooth non-periodic hazards, log-normal provided the best AIC and comparatively good PFS VPC performance.

Mechanistic MTLR avoided the need to select among these continuous baseline hazard families and showed the strongest descriptive PFS calibration of the parsimonious models in the clinical case study. Its model-specific PFS VPC contained the observed Kaplan-Meier curve at 93.4% of evaluation times, and its median simulated survival curve had the smallest mean absolute deviation from the observed Kaplan-Meier curve. The hybrid model achieved slightly higher VPC coverage, but its additional nonlinear weight was small and imprecisely estimated. This mirrors the simulated analysis, in which the hybrid extension provided localized gains in discrimination but did not consistently improve overall probabilistic accuracy. Across both datasets, the simpler mechanistic MTLR therefore captured most of the event-relevant information available from the longitudinal tumour trajectory. However, as a discrete-time survival approach, its performance can depend on how the time axis is discretized (Kvamme & Borgan, 2021). However, the results suggest that this flexibility is obtained without a major loss of predictive performance. In the simulated data analysis, Mechanistic MTLR had a slightly higher overall IBS than the best parametric hazard models, but achieved the lowest mean event-interval negative log score and superior discrimination at the later prediction landmarks. At 378–567 days, for example, mechanistic MTLR achieved an AUC of 0.867 compared with 0.798 for each of the conventional hazard formulations. These results indicate that the relative performance of MTLR depends on the aspect of prediction being evaluated. Conventional hazard models were somewhat stronger in overall probabilistic calibration, whereas MTLR was stronger in distinguishing individuals according to later event risk and in allocating probability to the appropriate event-time intervals. Time-dependent AUC evaluates discrimination in the presence of censored event times, whereas the Brier score evaluates error in predicted survival probabilities (Gerds & Schumacher, 2006; Heagerty et al., 2000).

The positive SLD-event association was also robust across the clinical hazard models. On the centered log(SLD + 1) scale, a doubling of SLD + 1 was associated with an approximately 1.41- to 1.55-fold increase in instantaneous PFS event hazard. The corresponding MTLR coefficients were also positive, although their magnitude is defined on the MTLR interval-score scale and should not be interpreted as a hazard ratio. The consistency in direction across event formulations supports the qualitative prognostic role of current tumour burden while highlighting that hazard and MTLR parameters operate on different mathematical scales. In the simulation study, the same consistency among the hazard models was seen with the identical AUCs obtained within each prediction window. Although changing the baseline hazard affected predicted probabilities and therefore Brier scores, it did not materially alter the ranking of individuals by event risk. This is consistent with the models sharing the same mechanistic SLD predictor and association structure. Hazard-function choice therefore appeared to influence calibration (predicted probability accuracy ) more strongly than discrimination in this simulated dataset. As with the clinical data analysis, MTLR, by contrast, changed both the representation of time and the mapping of the longitudinal state to event probabilities, which may explain its greater discrimination at later landmarks.

The later-landmark performance of mechanistic MTLR is particularly relevant to dynamic prediction. Joint longitudinal-survival models permit survival predictions to be updated as additional longitudinal measurements become available, thereby incorporating the evolving biomarker history into individualized risk estimates (Andrinopoulou et al., 2021; Rizopoulos, 2011; Rizopoulos et al., 2013). As additional longitudinal measurements became available, the patient- specific mechanistic SLD trajectory could be updated and incorporated into the interval-specific MTLR scores. The improved later AUC therefore demonstrates that an interval-based survival model can effectively exploit dynamically estimated longitudinal states without requiring those states to enter a prespecified continuous-time hazard function. This extends the main advantage of conventional MTLR to a joint pharmacometric setting in which the predictors themselves evolve through a nonlinear mixed-effects model.

The clinical longitudinal diagnostics reveal an important limitation. Individual observed-versus- predicted plots and residual summaries were satisfactory, but the population longitudinal VPC showed poor coverage of the 10th, median, and 90th SLD percentiles. Reducing the random- effects dimension solved the numerical identifiability problem but did not fully reproduce between-patient trajectory heterogeneity. The clinical event-model comparison should therefore be interpreted as conditional on a deliberately parsimonious longitudinal submodel. Future work should investigate whether one additional estimable dynamic random effect, for example on ϕ, or another parsimonious trajectory parameter improves population VPC performance without reintroducing the instability of the four-random-effect formulation.

The two analyses are not directly interchangeable. The simulated experiment used the original SLD/100 association and provided five-fold cross-validated prediction metrics, whereas the clinical analysis used log(SLD + 1) because of exact zero SLD values and heteroscedasticity. Treatment assignment and additional clinical covariates were intentionally excluded from the clinical case study, so its SLD-event associations should not be interpreted as treatment effects or as fully adjusted prognostic effects. In addition, the clinical VPCs were generated from the same dataset used for estimation and therefore assess model reproduction rather than independent predictive validation. The five-fold cross-validation from the simulated analysis remains the formal out-of-sample comparison. The simulation setting is both a strength and a limitation. It enabled comparison of event-model formulations without clinical heterogeneity, treatment changes, unmeasured confounding, or irregular observation processes obscuring the methodological differences. Additionally, with simulation studies the data-generating mechanism is known and factors such as sample size, censoring, and model misspecification can be systematically controlled (Morris et al., 2019). However, the findings should not be interpreted as evidence of clinical superiority or biological relevance of any particular model.

MTLR also replaces rather than eliminates structural modeling decisions. Its performance may depend on the number and placement of time intervals, regularization, and treatment of the final interval. Studies of discrete-time survival prediction have shown that discretization strategy can influence predictive performance, particularly in smaller datasets (Kvamme & Borgan, 2021). The modeling choice therefore shifts from selecting a continuous hazard family to selecting an appropriate discrete-time representation of the survival distribution. The combined simulated and clinical evidence suggests that this trade-off can be practically useful when baseline hazard shape is uncertain.

## Conclusion

Mechanistic MTLR provided a viable complementary alternative to conventional joint hazard models across both the controlled simulated experiment and the clinical PFS case study. In the simulated dataset, the best-performing parametric hazard model, particularly the log-normal formulation, showed slightly better overall probabilistic accuracy, whereas mechanistic MTLR achieved superior later-landmark discrimination and the best event-interval log score without requiring selection of a baseline hazard family. In the clinical dataset, mechanistic MTLR was numerically stable and reproduced the observed PFS distribution well, while the periodic hazard demonstrated how a flexible baseline hazard can capture features of scheduled progression assessment. The results therefore support mechanistic MTLR as a complementary joint-modeling approach to hazard-based TTE models, particularly when the appropriate hazard function is uncertain.

## Data Availability

All data produced in the present work are contained in the manuscript

## Acknowledgments

None

